# Governor partisanship explains the adoption of statewide mask mandates in response to COVID-19

**DOI:** 10.1101/2020.08.31.20185371

**Authors:** Christopher Adolph, Kenya Amano, Bree Bang-Jensen, Nancy Fullman, Beatrice Magistro, Grace Reinke, John Wilkerson

**Affiliations:** Department of Political Science, University of Washington, Seattle; Department of Health Metrics Sciences, University of Washington, Seattle

**Keywords:** masks, COVID-19, U.S. states, public policy, partisanship

## Abstract

Public mask use has emerged as a key tool in response to COVID-19. We develop and document a classification of statewide mask mandates that reveals variation in their scope and timing. Some U.S. states quickly mandated the wearing of face coverings in most public spaces, whereas others issued narrow mandates or no mandate at all. We consider how differences in COVID-19 epidemiological indicators and partisan politics affect when states adopted broad mask mandates, starting with the earliest broad public mask mandates in April 2020 and continuing though the end of 2020. The most important predictor is whether a state is led by a Republican governor. These states adopt statewide indoor mask mandates an estimated 98.0 days slower (95% CI: 88.8 to 107.3), if they did so at all (hazard ratio=7.54, 95% CI: 2.87 to 16.19). COVID-19 indicators such as confirmed cases or deaths per million are much less important predictors of statewide mask mandates. This finding highlights a key challenge to public efforts to increase mask-wearing, one of the most effective tools for preventing the spread of SARS-CoV-2 while restoring economic activity.

## Introduction

Public mask wearing is now widely viewed as a low-cost and effective means for reducing SARS-CoV-2 virus transmission (Chu, Akl, and Duda et al, 2020; Lyu and Wehby, 2020; Howard, Huang, and Li et al, 2020). However, it was not until 3 April 2020, more than a month after the first reported case of the novel coronavirus in the US, that the CDC formally recommended mask wearing to the general public (Centers for Disease Control and Prevention, 2020). Across the U.S., voluntary adherence to the CDC’s mask recommendation has been uneven. Mask wearing in response to airborne diseases is an established cultural norm in some societies, but this was not the case in the U.S. (Friedman, 2020).

As with other non-pharmaceutical interventions (NPIs) such as business and school closures and stay-at-home directives, (Adolph, Amano, Bang-Jensen, Fullman, and Wilkerson, 2021), the absence of a national mask mandate has led to considerable mask policy variation across states (Masks4All, 2020). Many U.S. states were slow to require citizens to wear masks across a broad range of indoor public spaces statewide, despite the CDC’s recommendation and despite growing evidence that mask wearing is an effective intervention. Only 33 of 50 states had adopted such mask mandates by the time United States’ second wave of COVID-19 began to recede in early August 2020. And amid the third wave of COVID-19 in the U.S., twelve states still lacked mandates by 31 December 2020 (Fullman et al 2021).^1^

Political scientists and democratic theorists assume that retrospective voting plays an important role in incentivizing good governance and accountability in democracies (Ashworth, 2012; Healy and Malhotra, 2013; Fearon, 1999; Ferejohn, 1986; Hopkins and Pettingill, 2018). The logic is that incumbents want to stay in office and fear that voters will throw them out if socioeconomic circumstances worsen during their terms. de Benedictis-Kessner and Warshaw (2020*a*), for instance, uncover evidence of retrospective voting even at the most local levels of government. If wages increase in a given state or county, it is the president’s party that is held responsible, an effect that only grows under conditions of heightened partisanship (de Benedictis-Kessner and Warshaw, 2020*b*).

By this logic, mandating masks would seem to be smart politics. Mask wearing reduces the risk of virus transmission, allowing people interact more safely among each other, and thus enabling faster and more sustained economic reopening. Indeed, other studies find that the pressures of fiscal federalism and the threat of exit by residents and businesses encourage state, local, and city politicians to pursue policies endorsed by experts rather than their national party bases (Peterson, 1981; Oates, 1999; Harmes, 2019).

Other research, however, provides reasons to question whether the conditions for retrospective voting are always present at the state level. The decline of journalism covering state and local politics, increasing polarization at the national level, and voters’ general inattention to down-ballot politics (Anzia, 2011; Rogers, 2016; Hopkins, 2018) may amplify the power of special interest groups, which likely gives state and local politicians more leeway in pursuing policies at odds with the well-being of residents. But what political incentives might lead state politicians to resist mask mandates? The possibility that special interests stand to lose if states adopt mask wearing policies seems implausible. A more likely explanation is partisan politics.

State politics are becoming increasingly nationalized and thus increasingly polarized along party lines (Shor and McCarty, 2011). Whether party polarization starts with voters, interest groups, or politicians themselves, the effect is greater polarization of state policy outcomes. This is a new development. Historically, studies found minimal partisan differences in state policy (Caughey, Warshaw, and Xu, 2017; Erikson, Wright, and McIver, 1993; Garand, 1988; Jacobs and Carmichael, 2002; Konisky, 2007). More recently, there is increasing evidence that party control is producing important policy differences across diverse policy areas (Adolph, Breunig, and Koski, 2020; Grumbach, 2018).

At first glance, variation in state mask mandate adoption appears to fall sharply along political party lines. Every one of the twelve states without a broad mask mandate at the end of 2020 was led by a Republican governor, and most of the early-adopting states were led by Democratic governors. But it is also possible that first impressions overlook the impact of other differences among states. For example, perhaps some states were slow to adopt or never adopted mask mandates because they had substantially fewer COVID-19 cases or deaths per capita.

Considering the mounting evidence that masks are an effective means for slowing the spread of SARS-CoV-2, and the rapid real-time policy innovation of mask mandates across states, it is important to uncover the central drivers of these state-level COVID-19 responses. By understanding these decisions, we may be able to better chart the landscape of future policy innovation in response to crisis.

Using originally collected data on mask mandates across states, we examine how variation in COVID-19 epidemiological indicators by state, partisan control of the executive, and other state characteristics may have affected the timing of mask mandate adoption. Specifically, we recorded when states issued, expanded, or rescinded mask mandates and developed a three-point scale to classify the breadth of each mandate. We then performed an event history analysis to explore variation in the timing of adoption of broad mandates that require individuals to wear masks while indoors in public spaces.

Controlling for state citizen ideology and the seven-day moving average of reported COVID-19 deaths per million residents, we find the governor’s party affiliation is the most important predictor of state differences in the timing of indoor public mask mandates. Over the nine-month span from 1 April 2020 to 31 December 2020, the marginal effect of a having Republican governor instead of a Democrat was a 98.0 day delay (95% CI: 88.8 to 107.3) in the issuance of broad state-wide mask mandates (hazard ratio=7.54, 95% CI: 2.87 to 16.19). This impact is far larger than that of any other variable examined and is robust to many different sensitivity analyses testing a large number of possible confounders and alternative measurements.

Why partisan politics is such a strong predictor of state mask mandates is beyond the scope of our analysis. We speculate that it is one more symptom of the nationalization of state level partisan politics and troubling evidence of the decline of retrospective policy voting. Republican governors feared being held to account by Republican voters, but not because they promoted policies that were detrimental to those voters’ health and economic well-being. Indeed, there is evidence that governors’ lack of action imposed considerable harm (Guy, Lee, and Sunshine et al, 2021).

Instead, the retrospective voting behavior these governors feared was being held to account for not supporting their partisan team. Opposing mask-wearing became a litmus test for Republicans. From the early stages of the pandemic, President Trump mocked mask-wearing as evidence of personal weakness, ignoring the role of masks in protecting other people from transmission of SARS-CoV-2 (Kaplan and Thrush, 2020). Republican governors therefore perceived that instructing their citizens to behave otherwise was politically perilous, despite the potential for prolonged health and economic costs for their state’s citizens. Although Republican lawmakers no longer risk direct backlash from the Trump administration for pursuing mask policies, antipathy toward masks among Republican party elites and Trump voters remains. Our analysis shows that the effects of the former president’s anti-mask stance have outlasted his tenure, and could be an enduring hallmark of the Republican platform.

## Data

We collected data on all statewide directives mandating masks issued from the start of the US epidemic through 31 December 2020. We consider a public mask mandate to be any policy that requires individuals to wear masks or other mouth and nose coverings when they are outside their places of residence. We include only mandates which apply to all individuals within a given setting, allowing exceptions for individuals with certain medical conditions or for young children. Our data thus do not include mandates which only require the use of masks or other personal protective equipment by employees (but not customers) as part of specific business operations.

To further capture variation across mask mandates applying to the general public, we create a typology with three ordered categories that encompass all statewide public mask mandates issued over this period:

### Limited mandate (Level 1)

Policies in this category involve limited mask mandates applying only to specific public settings. For example, mask mandates at this level might apply only to transportation services (e.g., issued by Vermont on May 1, augmented to a Level 3 policy on 24 July 2020 [State of Vermont, 2020*a,b*]), to retail establishments (e.g., issued by Alaska on 22 April 2020 and ended on 22 May 2020 [State of Alaska, 2020]), or to large gatherings where social distancing is not possible (e.g., issued by New Hampshire on 11 August 2020 [State of New Hampshire, 2020]). A common example of a limited mandate is one which applies only to people visiting state government buildings (e.g., issued by Utah on 26 June 26 2020 and Oklahoma on 16 November 2020 [State of Oklahoma, 2020; State of Utah, 2020]).

### Broad indoor mandate (Level 2)

Policies in this category constitute broad mask mandates requiring the use of masks or cloth face coverings by the public across most or all sectors of public activity indoors or in enclosed spaces. Mandates in this category may also include requirements that members of the public wear masks while waiting in line to enter an indoor space, or while using or waiting for shared transportation.

For example, Minnesota’s mask mandate (issued 22 July 2020) requires people over five years of age who are medically able to wear facial coverings or masks “in an indoor business or public indoor space, including when waiting outdoors to enter an indoor business or public indoor space, and when riding on public transportation, in a taxi, in a ride-sharing vehicle, or in a vehicle that is being used for business purposes” (State of Minnesota, 2020).

### Broad indoor and outdoor mandate (Level 3)

Policies in this most comprehensive category mandate the use of face coverings by the public across all public indoor spaces and in outdoor settings, though exceptions may be made for outdoor mask wearing where social distancing is possible. For instance, New York issued a mask policy on 15 April 2020 mandating all individuals who are medically able and over two years of age to wear a mask when in a public place and unable to maintain social distancing (State of New York, 2020). Washington state’s mask mandate, issued 24 June 2020, requires that “every person… wear a face covering that covers their nose and mouth when in any indoor or outdoor setting” (State of Washington, 2020).

Because Level 1 reflects a very limited mask mandate from the perspective of preventing transmission of the novel coronavirus, and because the policies within that category vary considerably from one another, we concentrate our analysis on adoption of mandates at Level 2 or 3: mandates that at a minimum include a broad requirement to wear masks indoors in public spaces. For these policies, we coded both the dates on which statewide policies were issued in each state at each level, as well as the date of enactment of each policy. Because our objective is to better understand the factors that influenced Governors’ decisions to implement mask mandates, we focus on the dates the policies were issued.

The top panel of Figure 1 shows when broad statewide mandates requiring masks in indoor public spaces (Level 2 or higher mandates) were adopted across the US, starting in April 2020. The bottom panel shows when the broadest dual indoor-outdoor mandates were adopted (Level 3). These adoptions occurred in three phases: from the middle of April to the end of May 2020, eleven states adopted Level 2 or higher mandates; most (eight states) were Level 3 mandates. The second phase began in mid-to late-June, and continued into early August. In this later phase, an additional 22 states adopted mask mandates of at least Level 2 or higher, bringing the total number of states with broad mandates to 33. Most of these (17) were also Level 3 mandates (for a total of 25 Level 3 mandates). Four of these 17 states (Maryland, Michigan, New Jersey, and Oregon) had already adopted Level 2 mandates in April. On 30 September 2020, Missis-sippi ended its statewide mask mandate, replacing it with county-level mask mandates which were coordinated by the state government.^2^ All other states adopting statewide mandates in the spring or summer maintained them through the end of 2020. The third phase of mask mandates began as US coronavirus cases increased in late fall: in November 2020, governors in North Dakota, Utah, Hawaii, Iowa, and New Hampshire introduced broad, statewide mask mandates, followed by Wyoming in December. In the same month, Virginia raised its mandate from Level 2 to Level 3. Thus, by the end of 2020, more than three-quarters of states, containing at least 78.7% of the U.S. population, had statewide mask mandates requiring masks in indoor settings. Sixty percent of states, containing 69.9% of the population, further required masks to be worn outdoors statewide.^3^

**Figure 1.**
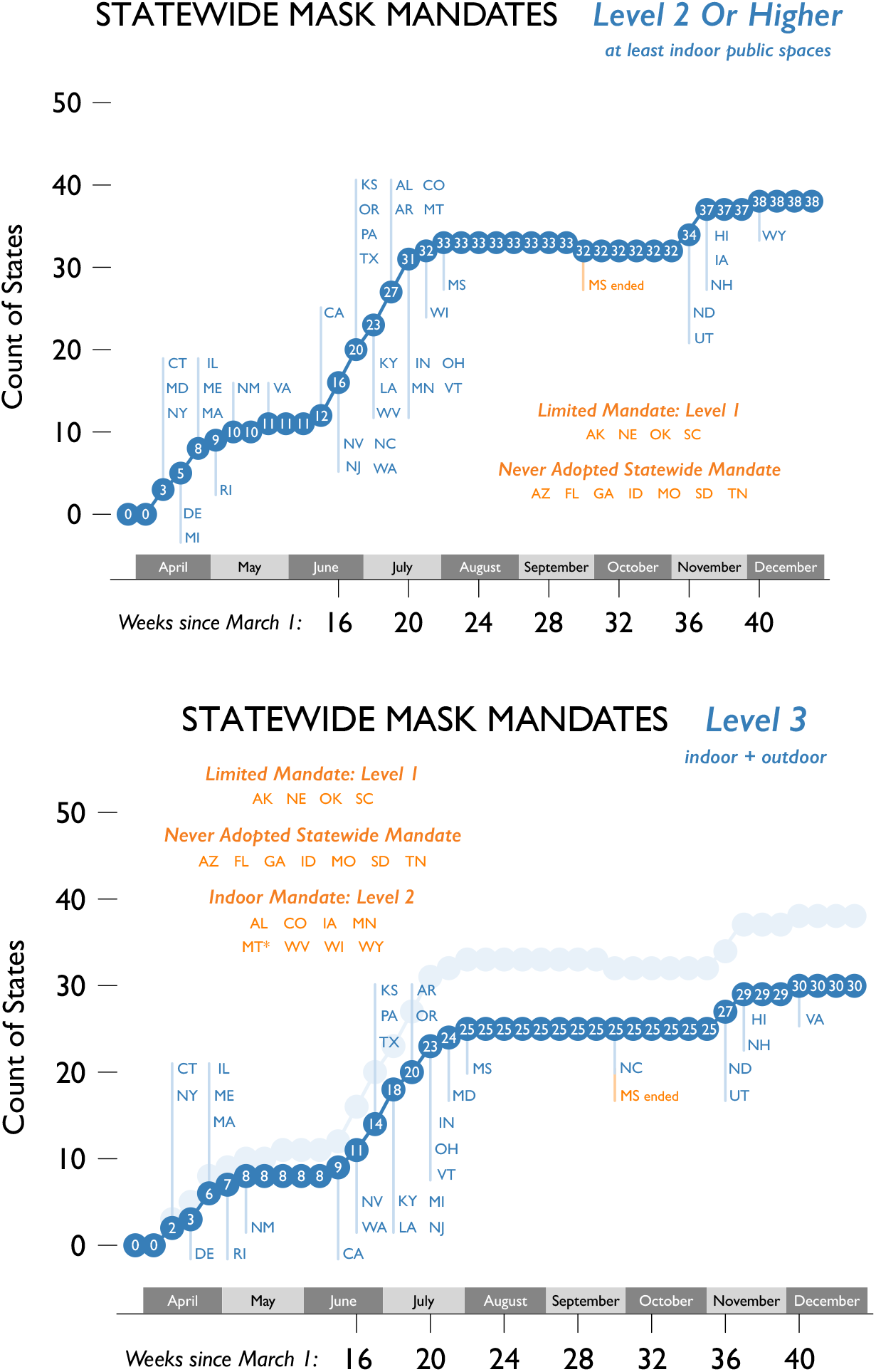
Adoption of broad statewide mask mandates in 2020. Weeks counted from 1 March 2020 for convenience. States listed as having Level 1 mandates are those which never adopted stronger mandates. Except for Mississippi, all states that introduced Level 2 or Level 3 statewide mask mandates maintained them at least through 31 December 2020. Except for Montana (which issued a Level 3 mandate on 13 January 2021, then repealed its mandate on 12 February 2021), no state adopted a higher statewide mask mandate in the first two months of 2021. Sources: Authors’ original data (Fullman et al 2021).

## Results

We use Cox proportional hazards models to explore how different factors influenced the timing of broad statewide mask mandates across the fifty U.S. states. These factors include COVID-19 indicators, state capacity, and partisan politics. Figure 2 reports the results from our baseline model, which controls for the log of the seven-day moving average of COVID-19 deaths per million population reported in the state, the ideological orientation of each state’s citizenry, and the party of the governor (New York Times, 2020; Fording, 2018; The National Conference of State Legislators, 2020). These results are reported both using traditional hazard ratios (top panel of Figure 2) and as average marginal effects across all fifty states, expressed as the average expected days of delay associated with each factor (bottom panel of Figure 2). The Methods Appendix provides further details of estimation and complete tables of results.

**Figure 2.**
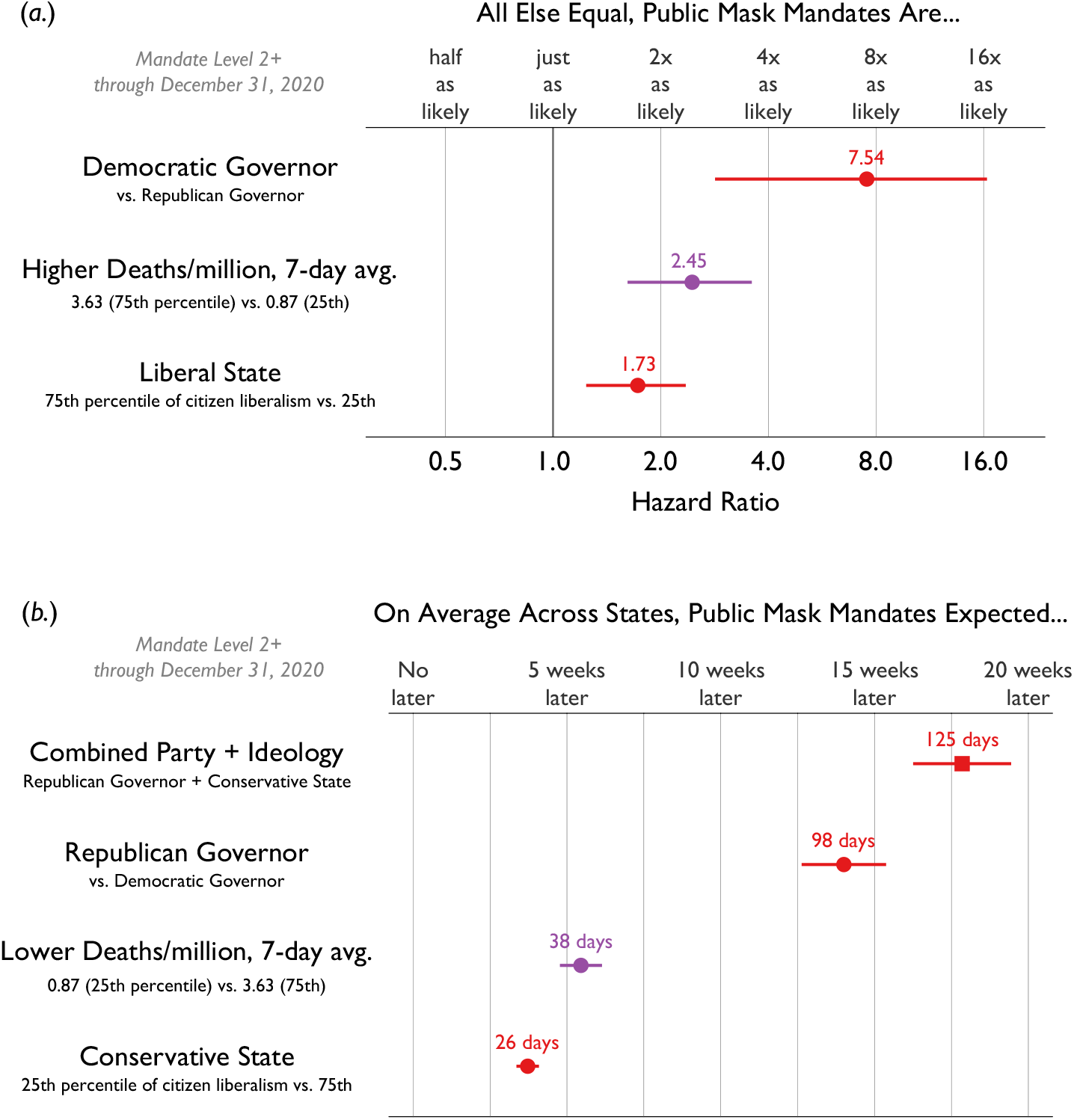
Relative probability (a.) and expected delay (b.) of adopting at least a Level 2 mask mandate, by factor. The top panel shows on a log scale the estimated hazard ratios obtained from a Cox proportional hazards model on mask mandates adopted by the fifty states, 1 April – 31 December 2020. Red circles mark the hazard ratios for political covariates, and purple circles indicate hazard ratios for other covariates. The bottom panel shows on a linear scale the estimated average marginal effects obtained by post-estimation simulation from the model. The red square marks combined effect of partisanship and ideology, red circles indicate the independent effects of governor party and citizen ideology, and purple circles indicate average marginal effects for other covariates. Horizontal lines are 95% confidence intervals. Solid symbols indicate significance at the 0.05 level.

By far, the most powerful predictor of broad mask mandate adoption and timing is the political party of the governor. Holding constant state ideology and the daily rate of COVID-19 deaths per million population, at any given time Democratic governors are 7.54 times more likely (95% CI: 2.87 to 16.19) to adopt a mask mandate of at least Level 2 than are Republican governors. We also use the estimated Cox model to predict the total expected delay hypothetically associated with having a Republican governor (rather than a Democratic governor) in each state, while leaving state ideology and COVID-19 deaths per million at their observed values for each state-day. We find that averaged across the fifty states, the marginal effect of having a Republican governor is a 98.0 day delay in adopting a broad indoor mask mandate (95% CI: 88.8 to 107.3 days).

The party of the governor is not the only political variable that influences the likelihood of adoption. Holding constant the party of the governor, states with more liberal citizens adopt mandates earlier than states with more conservative citizens. For example, states at the 75th percentile of citizen ideology (more liberal) are 1.73 times more likely to adopt mask mandates at a given time than more conservative states at the 25th percentile of citizen ideology (95% CI: 1.25 to 2.33). The marginal effect of this interquartile difference in citizen ideology is a 26.1 day delay of indoor mask mandates in more conservative states (95% CI: 23.9 to 28.3 days).

Researchers and policy-makers use several metrics to track SARS-CoV-2 transmission, and governors had access to daily data on COVID-19 measures including confirmed cases, deaths, and positive test result rates from both internal groups and state agencies. In our model, which uses epidemiological data from the New York Times, daily deaths per million dominates measures of new cases per million and test positivity rates (which we include among our robustness checks) as a factor associated with the timing of broad statewide mask mandates. Nevertheless, the effect of daily deaths is much weaker than the effect of governors’ party affiliation. We find that a state at the 75th percentile for daily COVID-19 deaths per million population is 2.45 times more likely to adopt a mask mandate at a given time than a state at the 25th percentile (95% CI: 1.63 to 3.56). Our model suggests a state with a persistently lower rate of daily COVID-19 deaths will adopt mask mandates 38.2 days later than a state with a higher daily death rate (95% CI: 33.7 to 42.6 days).

As Republican governors and conservative citizens often go together, the aggregate impact of politics on mask mandate adoption is even greater. When combined, the expected delay in adopting at least an indoor mask mandate for a state with both a Republican governor and a conservative citizenry is 124.9 days (95% CI: 114.1 to 135.7 days) when compared to a Democratic governor in a liberal state. The majority of this delay is attributable to the party of the executive, highlighting the importance of state-level political leadership in fighting the virus.

We conducted several additional analyses to test the robustness of these findings. First, we considered the possibility that our results were sensitive to either the source of daily COVID-19 data used in the model or the set of COVID-19 indicators used for each state-day. Our baseline model used data reported by the New York Times on daily COVID-19 deaths for each state (New York Times, 2020). Figure 3 reports results from a series of models that use alternative sources of daily death counts (The COVID Tracking Project, 2020; Center for Systems Science and Engineering, Johns Hopkins University, 2020). As the top of Figure 3 makes clear, the gap between the effect of governor partisanship and the effect of deaths per million remains at least as large as in the baseline model across the alternative indicators. Alternatively, instead of looking at the average rate of daily deaths, policymakers could be focused on trends.^4^ When added as an additional control to a model that already captures the rate of deaths, the trend in deaths has only a small effect on the risk of adopting a Level 2 or higher mask mandate, and the statistical significance of that effect is inconsistent across different data sources. Once again, the effect of partisan governors remains unchanged.

**Figure 3.**
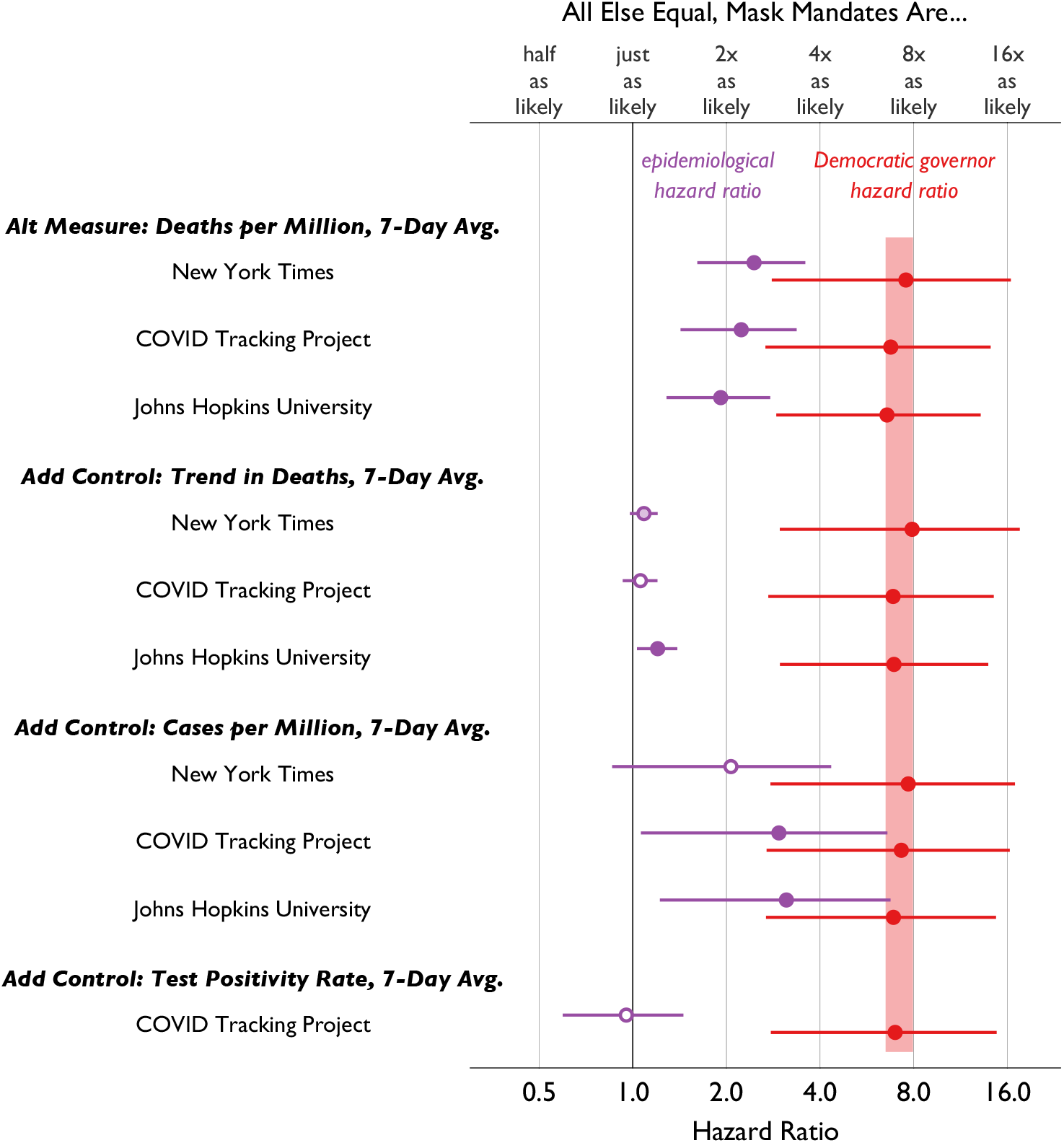
Sensitivity of results to alternative COVID-19 epidemiological indicators. Estimated hazard ratios of mask mandate adoption (Level 2 or higher) for various epidemiological indicators (in purple) and for Democratic governors (in red) from a series of Cox proportional hazards models adding each epidemiological covariate using data from the source listed at the left of the plot. Horizontal lines are 95% confidence intervals. Solid symbols indicate significance at the 0.05 level; shaded symbols indicate significance at the 0.1 level. Axes are log scaled.

COVID-19 deaths may be the most politically salient consequence of the pandemic, but they are also the least timely indicator of the severity of SARS-CoV-2 transmission in a given place and time, operating at a lag of approximately two or more weeks from the time of infection (Testa, Krieger, Chen, and Hanage, 2020; Wilson, Kvalsvig, Telfar Barnard, and Baker, 2020). We therefore consider models adding controls for more timely indicators of the spread of SARS-CoV-2: the number of confirmed COVID-19 cases per million reported in each state each day and the rate of test positivity (in both cases, as seven-day moving averages). Arguably, states taking prompt action to curb the spread of the virus should be responsive to these indicators. With respect to rates of confirmed cases, results are mixed and depend on the source of data. In a model using death and case data from the New York Times, the effect of higher rates of case growth is in the expected direction of encouraging mask mandates, but not statistically significant in a model that controls for the count of deaths. However, using data from either the COVID Tracking Project or Johns Hopkins University, we find significant relationships between confirmed cases per million population and adopting broad indoor mask mandates. On the other hand, rate of positive tests in a state had no relationship with mandate timing once deaths per million is controlled.^5^ In all models, the partisan effect was unchanged, and considerably larger than the effects of any epidemiological indicators.

Aside from alternative measures of public health indicators, we also consider a series of additional control variables, none of which alter our findings regarding the effect of partisan governors (Figure 4). First, we add a third measure of partisan politics, either Trump’s vote share in the state in the 2016 presidential election or the percentage of people in the state who watch Fox News regularly (New York Times Staff, 2017; Simply Analytics, 2018). Neither helps explain mask mandate timing in models that also control for governor party and citizen ideology. This may indicate that direct effects of these factors cannot be isolated, or that their impact on timing is mediated through governors and through their conservative audiences.

**Figure 4.**
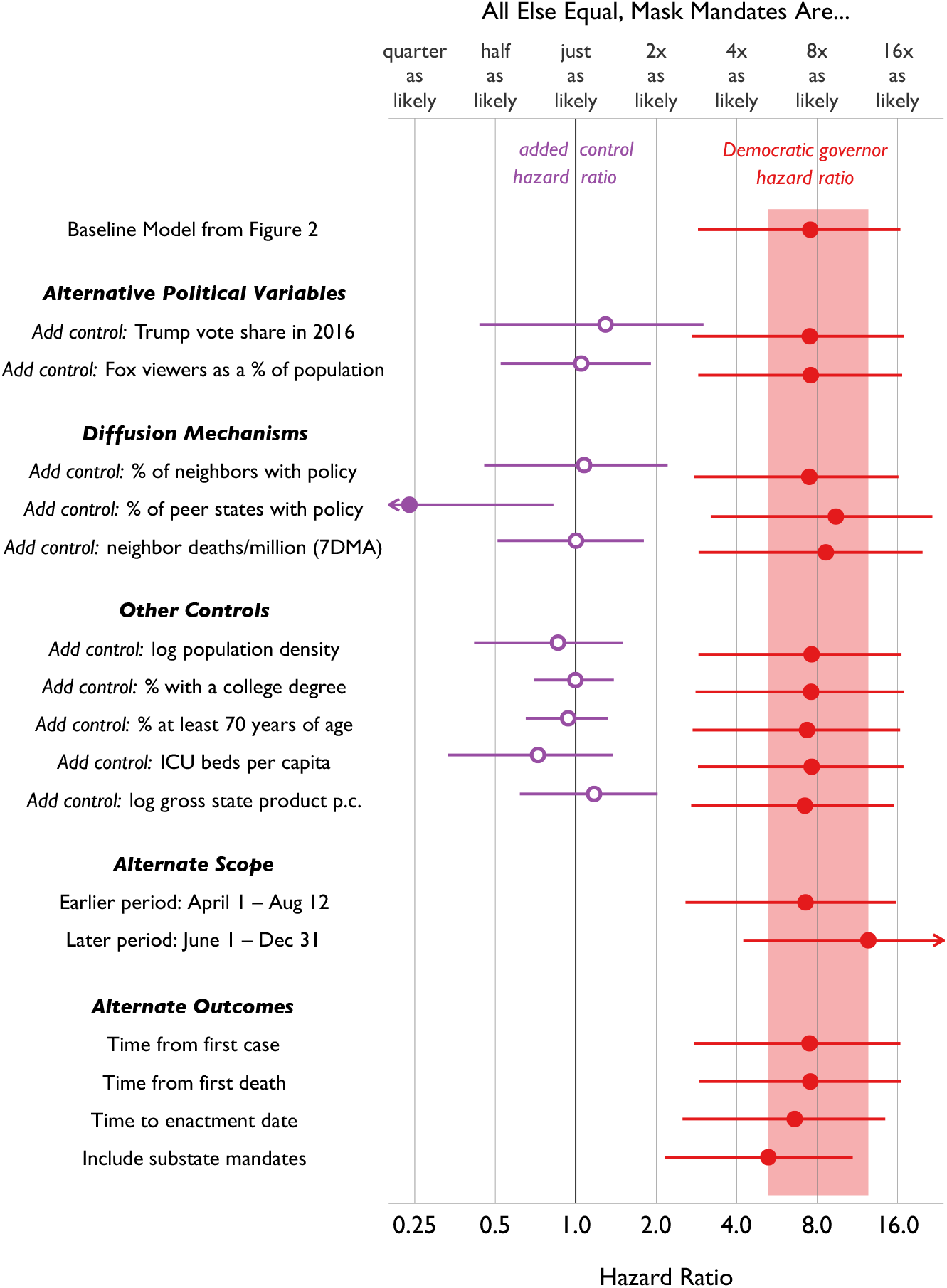
Democratic governors’ greater propensity to enact Mask Mandates is highly robust. Estimated hazard ratios of mask mandate adoption (Level 2 or higher) for effect of Democratic governors from a series of Cox proportional hazards models including various added controls or alternative outcome measures. Horizontal lines are 95% confidence intervals. Solid symbols indicate significance at the 0.05 level. Arrows indicate confidence intervals that extend outside the plotting range. Axes are log scaled. “Substate mandates” refers to mandates which apply to county-specific mandates coordinated by the state government and does not include local ordinances.

Next, we consider the possibility that states adopt (or fail to adopt) mask mandates either in imitation of policies adopted by other states, in reaction to the spread of the virus in neighboring states, or as an act of free-riding on the mandates of neighbors. We find that controlling for governor party, citizen ideology, and the daily death rate within a state, neither the adoption of mask mandates by neighboring states nor the average death rate in neighboring states is associated with the timing of mandates. We also control for the rate of mask mandate adoption in “peer states” – other states identified using network analysis as the innovators which a given state most often imitates across a variety of policy areas (Desmarais, Harden, and Boehmke, 2015). Because many peer states do not share a border, diffusion among peers is expected to be driven by patterns of policy imitation, rather than concern for spillovers or free-riding on neighbors’ restrictions. Puzzlingly, whether peer states have adopted mandates is negatively associated with mandate adoption once our baseline controls are included. We suspect this result is spurious, and in any case, inclusion of this control does not alter our main findings.

Other controls which fail to explain mandate timing when added to the model include the percentage of state residents above the age of 70 and the precentage of state residents in possession of a college degree (Institute for Health Metrics and Evaluation, 2017), as well as the log of population density (US Census, 2017) and the log of gross state product per capita (US Bureau of Economic Analysis, 2020). We consider the latter non-finding reasonable given the minimal economic consequences of a mask mandate, in contrast to many other non-pharmaceutical interventions. We consider one last control: the (pre-epidemic) count of ICU beds in each state per capita, which if low might add urgency to state policies to combat the pandemic (Harvard Global Health Institute, 2020). However, we find no significant association between pre-epidemic ICU beds and mask mandates, nor does inclusion of this control alter our main findings.

Next, we consider changes to the temporal scope of our analysis. In April and May 2020, states that adopted mask mandates did so either before they eased social distancing mandates, or concurrent with efforts to ease social distancing and re-open business sectors. Despite early ambivalence among medical experts about the effectiveness of masks as a COVID-19 NPI, these states may have issued mask mandates as a preventative policy layer to mitigate transmission risks associated with easing social distancing restrictions (Hendrix, Walde, Findley, and Trotman, 2020). Over time, the benefits of wearing non-medical masks against SARS-CoV-2 transmission were better understood and more widely publicized (Stutt, Retkute, Bradley, Gilligan, and Colvin, 2020; Javid and Balaban, 2020). Despite partisan resistance to mask mandates on the part of Republican voters and President Trump, one could imagine governors of both parties coalescing in June and July around mask mandates as the least costly intervention to protect fragile state economies and create a path to normal social interactions (Chu, Akl, and Duda et al, 2020; Lyu and Wehby, 2020).

Yet when we restrict our analysis to a later window of 1 June 2020 to 31 December 2020, we find an even stronger partisan governor effect (a hazard ratio of 11.00, 95% CI: 4.18 to 28.91) when compared to an earlier (albeit somewhat overlapping) window capturing just the first and second waves of the epidemic (from 1 April 2020 to 12 August 2020, the hazard ratio was 6.60, 95% CI: 2.70 to 16.16). These results confirm that the party of the governor was strongly associated with the adoption of mask mandates throughout 2020, rather than a short-lived phenomenon that could be explained by early reluctance to take the virus seriously in an uncertain, low-information environment. Instead, the context of an impending national election, and the sharp increase in partisan messaging that accompanied it, likely played a role in the persistence – and perhaps even intensification – of the partisan divide surrounding mask policies.

We also investigate alternative ways to measure the timing of mask mandates. In the preceding analyses, we considered states at risk of mask mandates starting from a common date (in most analyses, 1 April 2020), by which point the coronavirus had widely spread throughout the United States. An alternative is to “start the clock” for each state on the date the virus first showed up in that state as a confirmed case (or a confirmed death) to allow for the possibility that states which were slow to confirm the presence of the virus were biased against preventative action. Tailoring the set of state-days at risk to include only those days following either the first confirmed case or the first confirmed death in a given state does not change our results. Second, instead of measuring time to the issuance of mask mandates, we model the time to the enactment dates contained in those mandates. This change makes no substantive difference in our results. Third, the governors of three states – Mississippi, Ohio, and Oregon – imposed mask mandates of at least Level 2 for specific counties in advance of adopting statewide mandates.^6^ The preceding models ignore these early, geographically incomplete measures, but even if we assign adoption dates to these three states based on their earliest efforts to coordinate substate mask requirements, we obtain substantively similar results, suggesting our findings on partisan governors are not an artifact of focusing on statewide mandates.

As a final robustness check, we report a complementary analysis modeling the time to adoption of mandates requiring masks both indoors and outdoors (Level 3 mandates), a breadth of mandate only 60 percent of states adopted by the end of 2020. The results of this analysis are reported in Figure 5 as well as in Table 2 in the Methods Appendix. Overall, we find substantively similar results, with some quantitative differences from the size of hazard ratios associated with Level 3 and Level 2 or higher mandates, respectively. The hazard ratio associated with conservative citizen ideology grew to 2.54 in the model of Level 3 mandates (95% CI: from 1.77 to 3.56) from 1.73 in the more inclusive model of both Level 2 and Level 3 mandates, whereas the hazard ratios associated with the party of the governor and a higher moving-average of deaths per million each shrank somewhat. On a given day, adoption of Level 3 mask mandates were 2.54 times more likely under Democratic governors (95% CI: from 1.19 to 4.74), compared to 7.54 times for Level 2+ mandates. Finally, Level 3 mandates were 1.84 times more likely given a higher rate of deaths (95% CI: from 1.31 to 2.50), compared to 2.45 times for Level 2+ mandates. In all cases, these results remained significant at the 0.05 level and associated with substantively noteworthy average marginal effects. States with lower rates of daily deaths per million could be expected to adopt combined indoor-outdoor mask mandates 28.1 days later than states with low rates of daily deaths (95% CI: 24.8 to 31.4). The expected delay associated with Republican governors was 46.1 days (95% CI: 40.6 to 51.6), while more states with more conservative citizens could be expected to adopt Level 3 mandates 48.0 days later than states with liberal citizens (95% CI: 39.0 to 57.0). The combined delay for states with Republican governors and conservative citizens was 98.3 days (95% CI: 84.7 to 111.9).

**Figure 5.**
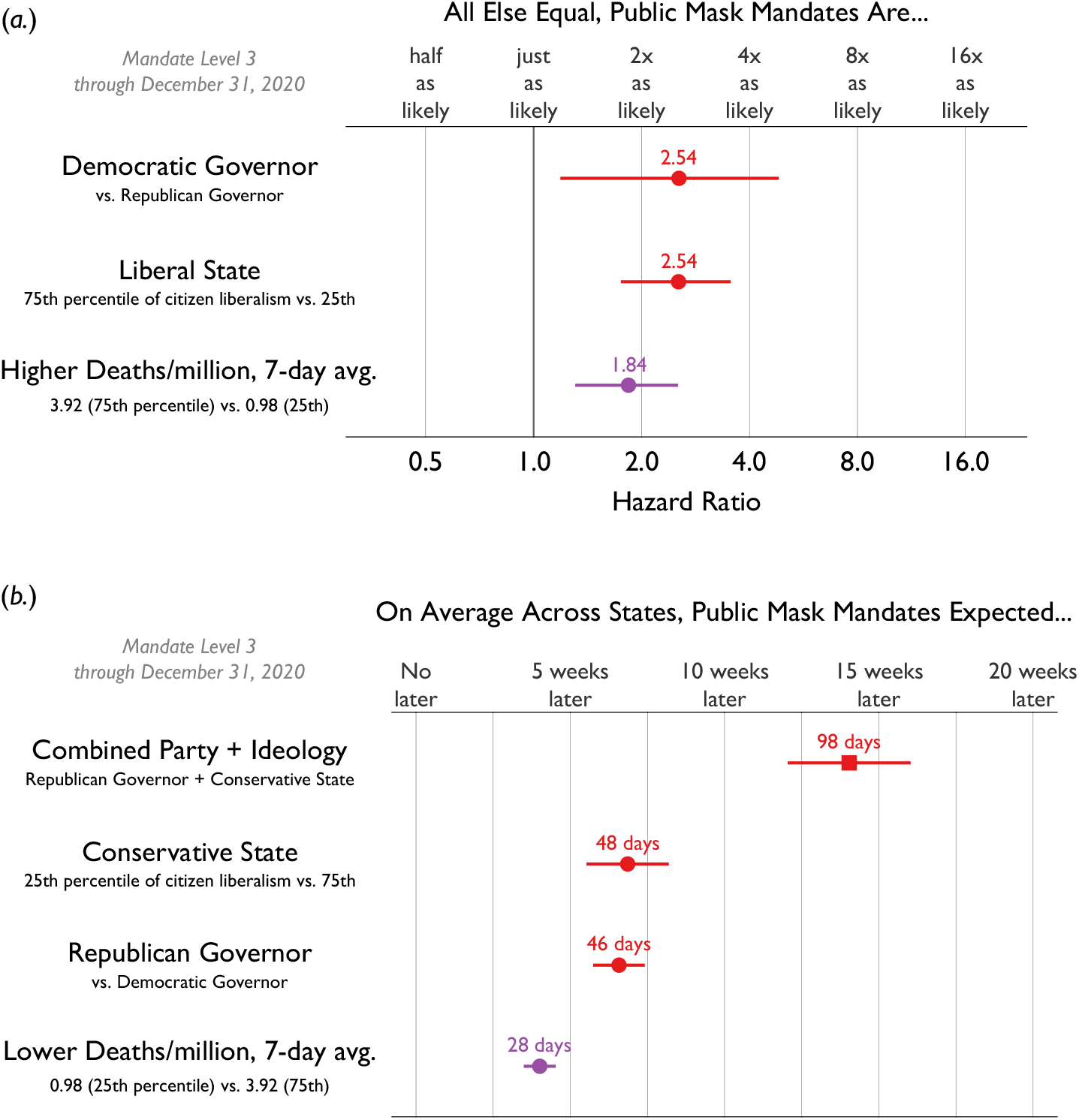
Relative probability (a.) and expected delay (b.) of adopting a Level 3 mandate for public masks, by factor. The top panel shows on a log scale the estimated hazard ratios obtained from a Cox proportional hazards model on mask mandates adopted by the fifty states, 1 April – 31 December 2020. Red circles mark the hazard ratios for political covariates, and purple circles indicate hazard ratios for other covariates. The bottom panel shows on a linear scale the estimated average marginal effects obtained by post-estimation simulation from the model. The red square marks combined effect of partisanship and ideology, red circles indicate the independent effects of governor party and citizen ideology, and purple circles indicate average marginal effects for other covariates. Horizontal lines are 95% confidence intervals. Solid symbols indicate significance at the 0.05 level.

## Discussion

Masks are an important, low-cost intervention to slow the spread of SARS-CoV-2 (Javid and Balaban, 2020; Stutt et al., 2020). Near-universal mask wearing reduces the risk implicit in returning to aspects of normal life and may be especially important for protecting essential workers who are not able to limit their exposure through social distancing (Javid and Balaban, 2020; Leatherby, 2020; Stutt et al., 2020).

In some countries, mask wearing is a well-established cultural norm (Friedman, 2020). This was not the case in the U.S. prior to COVID-19. By early summer 2020, governors of both parties surely recognized the pandemic’s continued threat to their states and were also well aware that it was transmitted via aerosols. One might therefore expect these leaders to be eager to encourage mask wearing as an alternative to the steep social and economic costs implicit in prolonged social distancing measures such as stay-at-home orders. The pandemic’s rapid progression also suggests that mandates, rather than public education campaigns, would be the preferred approach to ensuring mask compliance. Why, then, were so many Republican governors reluctant to promote this relatively low-cost and effective intervention?

Our event history analysis cannot pin down exact motives. The most likely explanation, we believe, is that the absence of a mask wearing norm in the U.S. opened the door to reactionary responses. From the beginning of the pandemic, President Trump seemed more concerned about the pandemic’s threat to the economy than the threat to public health. He and other Republican party elites may have seen mask wearing as a constant public reminder of a problem they were trying to minimize. The President publicly mocked mask wearers and continued to oppose a national mandate even after becoming seriously ill himself (BBC, 2020).

However, presidents need support from other party leaders, especially in public health, where the states have the greatest constitutional authority to act. Given the reality of the pandemic, why did the President’s position have so much support among other Republican leaders? The U.S. is as polarized politically as it has ever been, including across and within state governments (Grumbach, 2020; Masket, 2009; Shor and McCarty, 2011). A plausible hypothesis is that partisan politics motivated many Republican governors to oppose or delay imposing mask mandates, not because they truly believed masks to be ineffective or unnecessary, but because Trump’s very public rejection of mask wearing influenced the attitudes of a significant fraction of Republican voters (Gallup, 2020). In the midst of the epidemic, Republican identifiers were much less likely than Democratic identifiers to say that they wear masks all or most of the time (53% vs. 76% in August 2020 Igielnik, 2020). Republicans were also more likely to resist mask wearing as a sign of weakness or “unmanly” behavior (Capraro and Barcelo, 2020; Glick, Berdahl, and Alonso, 2018; Glick, 2020), perhaps based on the mistaken assumption that self-protection is the primary objective of mask wearing.

In this context, a Republican governor who mandated masks risked being portrayed as weak, threatening their base of support and possibly the support of their party’s national leader. Democratic governors, in contrast, had their own political reasons to support mask-wearing. These included: a generally positive view of mask wearing among Democratic constituents (Clinton, Cohen, Lapinski, and Trussler, 2020; Katz, Sanger-Katz, and Quealy, 2020; Igielnik, 2020); antipathy toward Trump, including his cavalier treatment of experts within his own government; and widespread mask wearing by other Democratic elites, including Joe Biden.

As cases and deaths rose in their states, some Republican governors like Kay Ivey of Alabama eventually accepted the need for mask mandates. But numerous other Republican governors, notably Kristi Noem in South Dakota, Brian Kemp in Georgia, Ron DeSantis in Florida, and Doug Ducey in Arizona resisted statewide mandates throughout even the third wave of the US epidemic. By the end of 2020, 38 states required masks indoors (Level 2 or higher). All twelve of the states that did not have broad indoor mandates at the end of the year were led by Republican governors.

In many localities, mask-wearing norms seem to have developed that will have positive longer term consequences for pandemic response (van der Westhuizen, Kotze, Tonkin-Crine, Gobat, and Greenhalgh, 2020). However, partisan politics delayed the adoption of an affordable and effective intervention to reduce coronavirus spread. There is reason to be concerned that this politicization of public health may also hinder future public health efforts that depend on rapid and widespread compliance, coherent elite messaging, and public buy-in.

In early 2021, with the third wave still ebbing, new variants of the coronavirus spreading in the United States, and vaccination rates still too low to prevent further surges, four states with Republican governors ended their statewide mandates: North Dakota (18 January 2021), Iowa (7 February 2021), Montana (12 February 2021), and Texas (10 March 2021). Although Trump is no longer in office, his enduring support among Republican voters means Republican leaders still have limited incentives to support ongoing mitigation efforts against COVID-19. In the short-run, this absence of leadership is of special concern given high levels of vaccine hesitancy among Republican self-identifiers. As of early March 2021, 41% of Republicans and 49% of Republican men surveyed would refuse a COVID-19 vaccine, compared to 30 percent of all Americans, 11% of Democrats, and just 6% of Democratic men (Marist Poll, 2021).

In the longer run, the finding that increasing party polarization blocked even the most cost-effective measures for combating COVID-19 bodes poorly for future responses to public health crises. Functioning representative democracies depend on accountability. Elected officials must believe that voters may hold them accountable for harmful socioeconomic outcomes. This retrospective voting model is threatened when voters are successfully encouraged to view issues largely in terms of us versus them. The tragedy, of course, is that infectious diseases do not make partisan distinctions, nor are they confined within state lines. The consequences of public policy failure are not confined to those who oppose public health interventions, but are suffered by all.

## Methods Appendix

We estimate an event history model to predict the timing of announced mask mandates across U.S. states from 1 April 2020 to 31 December 2020. Specifically, we model the likelihood that a state will implement a mask mandate of at least Level 2 (broadly requiring face coverings indoors) as a function of time in days with a Cox proportional hazards model, clustering standard errors by state. All states are considered at risk of adopting a mandate starting on April 1, and remain at risk until they adopt a mandate at either Level 2 or Level 3. In this model, the baseline hazard rate non-parametrically captures the effects of purely national trends – such as the common tendency of states to adopt mask mandates due to the national resurgence of new COVID-19 cases and deaths, or as a result of new scientific findings regarding the effectiveness of masks in reducing coronavirus transmission. This leaves only cross-state variation in the timing of mask mandates to be explained by covariates.

Our primary specification, reported in Table 1, includes two time-invariant covariates – the ideological orientation of each state’s citizenry and the party of the governor (Fording, 2018; The National Conference of State Legislators, 2020). We also control for a time-varying covariate, the daily reported COVID-19 deaths per million population in each state using case data from the New York Times (New York Times, 2020) and population data from the US Census (Institute for Health Metrics and Evaluation, 2017). Deaths per million enter the model both as a seven-day average, to smooth over differential rates of reporting over weekends and weekdays, and logged, to allow for diminishing marginal effects of rising COVID-19 deaths and to mitigate the influence of outliers, which in some cases likely reflect idiosyncratic reporting delays.

**Table 1.**
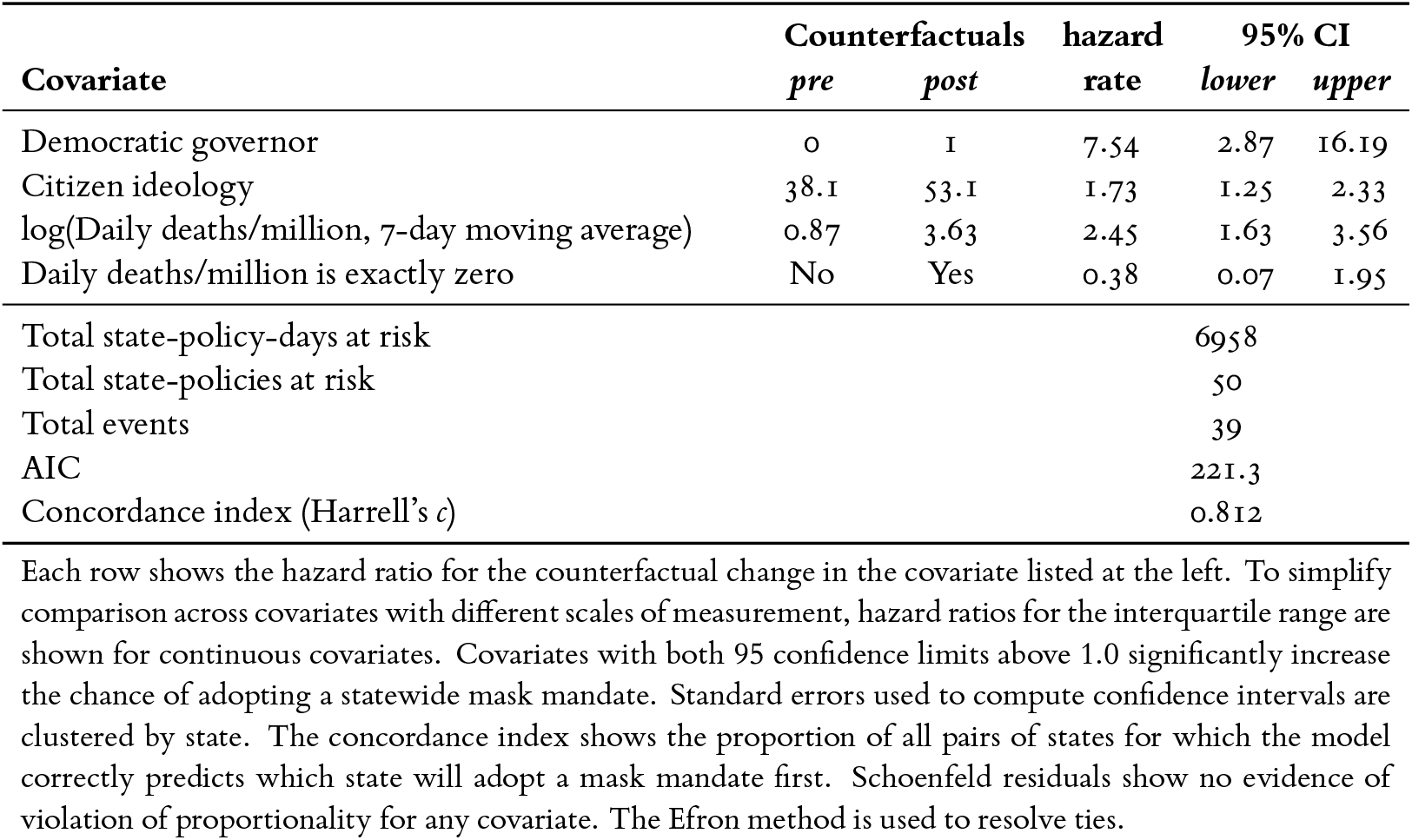
Hazard ratios from the baseline Cox proportional hazards model of state-level mask mandates, Level 2 or higher, 1 April to 31 December 2020.

| Covariate | Counterfactuals |  | hazard rate | 95% CI |  |
| --- | --- | --- | --- | --- | --- |
|  | *pre* | *post* |  | *lower* | *upper* |
| Democratic governor | 0 | 1 | 7.54 | 2.87 | 16.19 |
| Citizen ideology | 38.1 | 53.1 | 1.73 | 1.25 | 2.33 |
| log(Daily deaths/million, 7-day moving average) | 0.87 | 3.63 | 2.45 | 1.63 | 3.56 |
| Daily deaths/million is exactly zero | No | Yes | 0.38 | 0.07 | 1.95 |
| Total state-policy-days at risk |  |  |  | 6958 |  |
| Total state-policies at risk |  |  |  | 50 |  |
| Total events |  |  |  | 39 |  |
| AIC |  |  |  | 221.3 |  |
| Concordance index (Harrell's *c*) |  |  |  | 0.812 |  |

Each row shows the hazard ratio for the counterfactual change in the covariate listed at the left. To simplify comparison across covariates with different scales of measurement, hazard ratios for the interquartile range are shown for continuous covariates. Covariates with both 95 confidence limits above 1.0 significantly increase the chance of adopting a statewide mask mandate. Standard errors used to compute confidence intervals are clustered by state. The concordance index shows the proportion of all pairs of states for which the model correctly predicts which state will adopt a mask mandate first. Schoenfeld residuals show no evidence of violation of proportionality for any covariate. The Efron method is used to resolve ties.

Logging this term improves model fit (concordance increases from 0.809 to 0.812), but poses the problem of how to deal with seven-day averages over periods with no reported deaths. A common but flawed solution is to add a small “fudge” factor (e.g., 0.01, or 1, etc.) to cases of zero deaths to ensure the log of deaths per million is always defined; however, this technique produces different results depending on the (arbitrary) amount added. This is an underappreciated but unsurprising problem, as the range of plausible adjustments covers several orders of magnitude. While differences across plausible “fudge” factors do not affect our substantive or statistical conclusions enough to change our findings, a non-arbitrary solution is preferable. Instead, we rely on the data to suggest the appropriate treatment of zeros by including an additional covariate indicating cases where the moving average of deaths is exactly zero. In turn, before logging the moving average of deaths, we replace zeros with ones, ensuring (without loss of generality) that the zero cases “drop out” of the log term. The results from this zero-adjusted log specification are similar to those from models that use a “fudge” factor, but arguably less arbitrary and more data-driven.

We present our results in several formats. The exponentiated coefficients for every model we estimated (including all sensitivity analyses shown in Figures 3 and 4) are contained in Tables 3, 4, and 5 at the end of this appendix. However, because many of our covariates are continuous, exponentiated coefficients can be difficult to directly interpret or compare. Instead, hazard ratios for substantively relevant counterfactual scenarios associated with each covariate in our primary model are reported in Table 1. For continuous covariates, we show the hazard ratio associated with an interquartile shift in the covariate, as recommended by Harrell (2015). These are the hazard ratios reported in the top panel of Figure 2 in the main text. Following the approach of Adolph et al. (2021), we further contextualize these findings by computing the average marginal effect of each covariate averaged across the fifty states (Harden and Kropko, 2019), expressed as the expected days of delay associated with each covariate, averaged across the fifty states with all other covariates taking on their observed values day by day for each state. These quantities are shown in the bottom of Figure 2 in the main text. In parallel fashion, we collect in Table 2 the counterfactual hazard ratios for the model of Level 3 mandates shown in Figure 5 in the main text.

**Table 2.** Hazard ratios from the baseline Cox proportional hazards model of state-level mask mandates, Level 3 only, 1 April to 31 December 2020.

| Covariate | Counterfactuals |  | hazard rate | 95% CI |  |
| --- | --- | --- | --- | --- | --- |
|  | *pre* | *post* |  | *lower* | *upper* |
| Democratic governor | 0 | 1 | 2.54 | 1.19 | 4.74 |
| Citizen ideology | 38.2 | 53.8 | 2.54 | 1.77 | 3.56 |
| log(Daily deaths/million, 7-day moving average) | 0.98 | 3.92 | 1.84 | 1.31 | 2.50 |
| Daily deaths/million is exactly zero | No | Yes | 0.35 | 0.10 | 1.27 |
| Total state-policy-days at risk |  |  |  | 8531 |  |
| Total state-policies at risk |  |  |  | 50 |  |
| Total events |  |  |  | 31 |  |
| AIC |  |  |  | 193.9 |  |
| Concordance index (Harrell's *c*) |  |  |  | 0.792 |  |

Each row shows the hazard ratio for the counterfactual change in the covariate listed at the left. To simplify comparison across covariates with different scales of measurement, hazard ratios for the interquartile range are shown for continuous covariates. Covariates with both 95 confidence limits above 1.0 significantly increase the chance of adopting a statewide mask mandate. Standard errors used to compute confidence intervals are clustered by state. The concordance index shows the proportion of all pairs of states for which the model correctly predicts which state will adopt a mask mandate first. Schoenfeld residuals show no evidence of violation of proportionality for any covariate. The Efron method is used to resolve ties.

**Table 3.** Cox proportional hazards models of state-level mask mandates, Level 2 or higher: alternative epidemiological data.

| Covariates | 1 | 2 | 3 | 4 | 5 | 6 | 7 | 8 | 9 | 10 |
| --- | --- | --- | --- | --- | --- | --- | --- | --- | --- | --- |
| Democratic governor | 6.828<br><0.001 | 6.198<br><0.001 | 6.119<br><0.001 | 6.958<br><0.001 | 6.563<br><0.001 | 6.268<br><0.001 | 7.113<br><0.001 | 6.265<br><0.001 | 6.430<br><0.001 | 6.374<br><0.001 |
| Citizen ideology | 1.848<br><0.001 | 1.718<br><0.001 | 1.59<br>0.001 | 1.04<br><0.001 | 1.041<br><0.001 | 1.044<br>0.001 | 1.029<br>0.012 | 1.036<br>0.002 | 1.028<br>0.025 | 1.037<br>0.001 |
| log(Daily deaths/million,<br>7-day moving average) | 1.036<br>0.001 | 1.036<br>0.001 | 1.034<br>0.010 | 1.462<br>0.043 | 1.195<br>0.400 | 1.153<br>0.400 | 1.662<br>0.001 | 1.659<br>0.001 | 1.346<br>0.043 | 1.828<br>0.006 |
| Daily deaths/million<br>is exactly zero | 0.379<br>0.246 | 0.363<br>0.234 | 0.545<br>0.268 | 0.581<br>0.529 | 0.775<br>0.804 | 0.618<br>0.401 | 0.440<br>0.341 | 0.359<br>0.235 | 0.614<br>0.395 | 0.346<br>0.229 |
| log(Daily cases/million,<br>7-day moving average) |  |  |  | 1.461<br>0.117 | 1.769<br>0.037 | 1.839<br>0.015 |  |  |  |  |
| Linear trend in daily deaths<br>7-day moving average |  |  |  |  |  |  | 2.230<br>0.079 | 1.677<br>0.367 | 7.353<br>0.009 |  |
| Test positivity rate,<br>7-day moving average |  |  |  |  |  |  |  |  |  | 0.357<br>0.751 |
| Epidemiological data | NYT | CTP | JHU | NYT | CTP | JHU | NYT | CTP | JHU | CTP |
| State-policy-days at risk | 6958 | 6958 | 6958 | 6958 | 6958 | 6958 | 6958 | 6958 | 6958 | 6958 |
| Total state-policies at risk | 50 | 50 | 50 | 50 | 50 | 50 | 50 | 50 | 50 | 50 |
| Total events | 39 | 39 | 39 | 39 | 39 | 39 | 39 | 39 | 39 | 39 |
| AIC | 221.3 | 225.7 | 228.5 | 221.5 | 223.8 | 224 | 221.5 | 227.0 | 224.7 | 227.6 |
| Concordance index | 0.812 | 0.797 | 0.797 | 0.823 | 0.818 | 0.823 | 0.817 | 0.798 | 0.811 | 0.798 |

**Table 4.** Cox proportional hazards models of state-level mask mandates, Level 2 or higher: additional control variables.

| Covariates | 11 | 12 | 13 | 14 | 15 | 16 | 17 | 18 | 19 | 20 |
| --- | --- | --- | --- | --- | --- | --- | --- | --- | --- | --- |
| Democratic governor | 6.726<br><0.001 | 6.831<br><0.001 | 6.819<br><0.001 | 8.337<br><0.001 | 7.625<br><0.001 | 6.915<br><0.001 | 6.867<br><0.001 | 6.629<br><0.001 | 6.890<br><0.001 | 6.586<br><0.001 |
| Citizen ideology | 1.030<br>0.124 | 1.036<br>0.001 | 1.036<br>0.003 | 1.046<br><0.001 | 1.053<br><0.001 | 1.043<br>0.004 | 1.037<br>0.002 | 1.039<br>0.002 | 1.046<br>0.001 | 1.035<br>0.001 |
| log(Daily deaths/million,<br>7-day moving average) | 1.880<br><0.001 | 1.845<br><0.001 | 1.853<br>0.001 | 1.808<br><0.001 | 1.570<br>0.058 | 1.937<br><0.001 | 1.851<br><0.001 | 1.853<br><0.001 | 1.784<br><0.001 | 1.843<br><0.001 |
| Daily deaths/million<br>is exactly zero | 0.369<br>0.243 | 0.381<br>0.254 | 0.378<br>0.246 | 0.455<br>0.367 | 0.417<br>0.149 | 0.330<br>0.171 | 0.378<br>0.247 | 0.380<br>0.244 | 0.432<br>0.309 | 0.388<br>0.261 |
| Trump vote share 2016 | 0.989<br>0.769 |  |  |  |  |  |  |  |  |  |
| Fox viewers,<br>% of population |  | 1.003<br>0.975 |  |  |  |  |  |  |  |  |
| % of neighbors<br>mask mandate |  |  | 1.000<br>0.979 |  |  |  |  |  |  |  |
| % of peers<br>mask mandate |  |  |  | 0.964<br>0.028 |  |  |  |  |  |  |
| log(Mean neigh. deaths/m,<br>7-day moving average) |  |  |  |  | 0.953<br>0.870 |  |  |  |  |  |
| Neighbor daily deaths/m<br>is exactly zero |  |  |  |  | 0.099<br><0.001 |  |  |  |  |  |
| log(Population density) |  |  |  |  |  | 0.893<br>0.516 |  |  |  |  |
| % College degree<br>or higher |  |  |  |  |  |  | 0.996<br>0.910 |  |  |  |
| % 70 years or older |  |  |  |  |  |  |  | 0.926<br>0.645 |  |  |
| ICU beds per<br>10k population |  |  |  |  |  |  |  |  | 1.472<br>0.270 |  |
| log(Gross state product<br>per capita) |  |  |  |  |  |  |  |  |  | 1.565<br>0.705 |
| State-policy-days at risk | 6958 | 6958 | 6958 | 6958 | 6958 | 6958 | 6958 | 6958 | 6958 | 6958 |
| Total state-policies at risk | 50 | 50 | 50 | 50 | 50 | 50 | 50 | 50 | 50 | 50 |
| Total events | 39 | 39 | 39 | 39 | 39 | 39 | 39 | 39 | 39 | 39 |
| AIC | 223.2 | 223.3 | 223.3 | 220.6 | 220.0 | 222.9 | 223.3 | 223.1 | 222.1 | 223.2 |
| Concordance index | 0.811 | 0.814 | 0.813 | 0.823 | 0.819 | 0.811 | 0.815 | 0.820 | 0.812 | 0.815 |

**Table 5.**
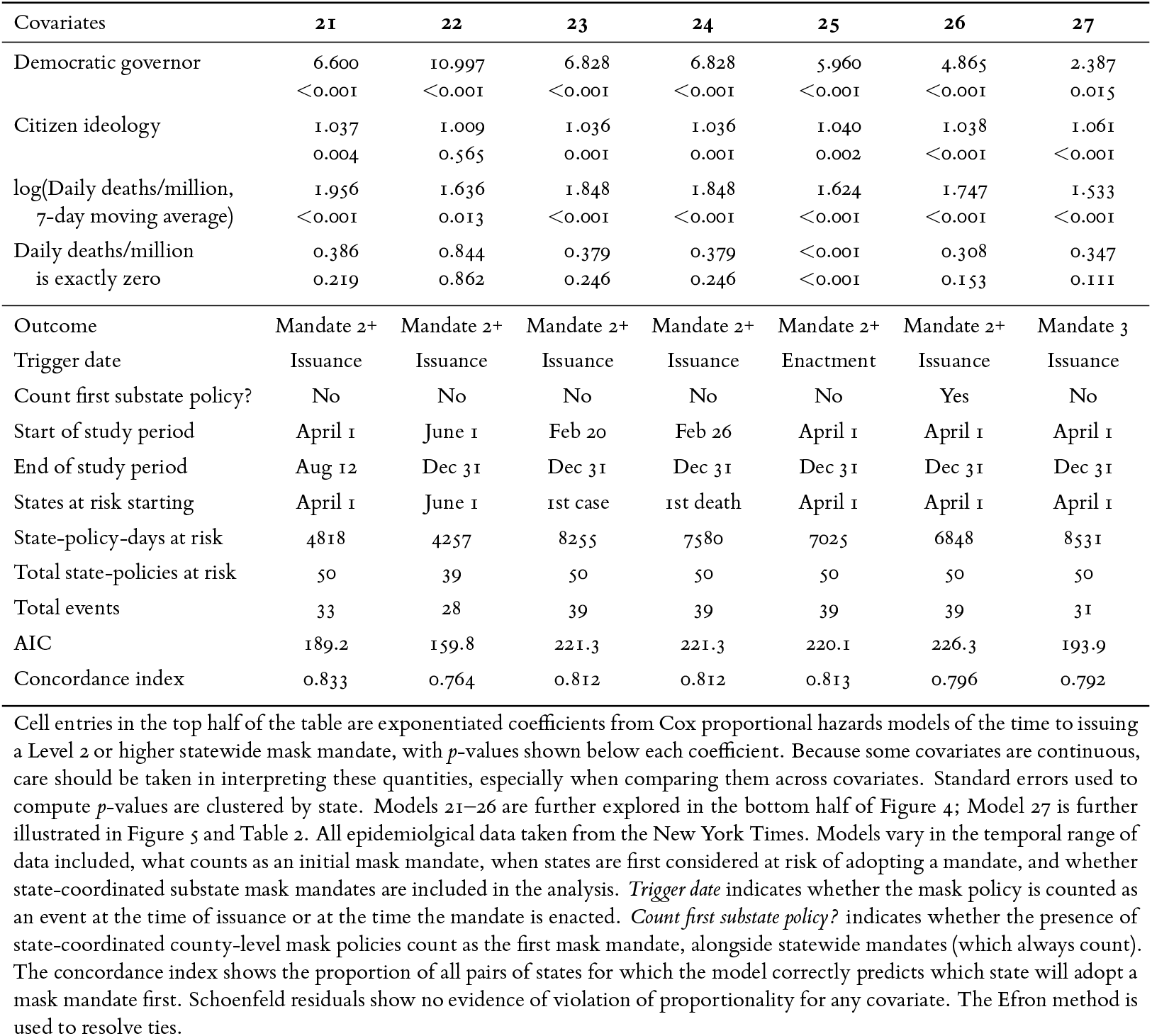
Cox proportional hazards models of state-level mask mandates: alternative scope conditions and outcome measures.

| Covariates | 21 | 22 | 23 | 24 | 25 | 26 | 27 |
| --- | --- | --- | --- | --- | --- | --- | --- |
| Democratic governor | 6.600<br><0.001 | 10.997<br><0.001 | 6.828<br><0.001 | 6.828<br><0.001 | 5.960<br><0.001 | 4.865<br><0.001 | 2.387<br>0.015 |
| Citizen ideology | 1.037<br>0.004 | 1.009<br>0.565 | 1.036<br>0.001 | 1.036<br>0.001 | 1.040<br>0.002 | 1.038<br><0.001 | 1.061<br><0.001 |
| log(Daily deaths/million,<br>7-day moving average) | 1.956<br><0.001 | 1.636<br>0.013 | 1.848<br><0.001 | 1.848<br><0.001 | 1.624<br><0.001 | 1.747<br><0.001 | 1.533<br><0.001 |
| Daily deaths/million<br>is exactly zero | 0.386<br>0.219 | 0.844<br>0.862 | 0.379<br>0.246 | 0.379<br>0.246 | <0.001<br><0.001 | 0.308<br>0.153 | 0.347<br>0.111 |
| Outcome | Mandate 2+ | Mandate 2+ | Mandate 2+ | Mandate 2+ | Mandate 2+ | Mandate 2+ | Mandate 3 |
| Trigger date | Issuance | Issuance | Issuance | Issuance | Enactment | Issuance | Issuance |
| Count first substate policy? | No | No | No | No | No | Yes | No |
| Start of study period | April 1 | June 1 | Feb 20 | Feb 26 | April 1 | April 1 | April 1 |
| End of study period | Aug 12 | Dec 31 | Dec 31 | Dec 31 | Dec 31 | Dec 31 | Dec 31 |
| States at risk starting | April 1 | June 1 | 1st case | 1st death | April 1 | April 1 | April 1 |
| State-policy-days at risk | 4818 | 4257 | 8255 | 7580 | 7025 | 6848 | 8531 |
| Total state-policies at risk | 50 | 39 | 50 | 50 | 50 | 50 | 50 |
| Total events | 33 | 28 | 39 | 39 | 39 | 39 | 31 |
| AIC | 189.2 | 159.8 | 221.3 | 221.3 | 220.1 | 226.3 | 193.9 |
| Concordance index | 0.833 | 0.764 | 0.812 | 0.812 | 0.813 | 0.796 | 0.792 |

In addition to the primary model reported in the Table 1 and Figure 2, we consider a series of sensitivity analyses of our model of Mandates at Level 2+. Throughout these analyses, we attempt to keep each estimated model parsimonious, as including too many covariates is a particular concern for event history models with small numbers of observed events (Peduzzi, Concato, Feinstein, and Holford, 1995). The first sensitivity analyses reported in Figure 3 simply replace the New York Times death data used in the primary model with data from alternative sources (The COVID Tracking Project, 2020; Center for Systems Science and Engineering, Johns Hopkins University, 2020). However, most of the sensitivity analyses, including those shown in the top half of Figure 3 retain the covariates of the primary model and serially add a single additional covariate. The underlying estimates for each sensitivity analysis are collected in Tables 3, 4, and 5.

Analyses were performed in R (version 4.0.2) using the survival and coxed (Harden and Kropko, 2019) packages. Across every model presented in the paper, the Schoenfeld residuals for each covariate shows no evidence of violation of proportionality, supporting the proportional hazard assumption. All visualizations were constructed using the title package (Adolph, 2020).

## Data Availability

State mask policy data are available at http://covid19statepolicy.org.

## Acknowledgements

The authors thank Rachel Castellano, Carolyn Dapper, Megan Erickson, and Rebecca Walcott for policy coding assistance, Erika Steiskal for graphic design assistance, and Jake Grumbach for helpful suggestions. We gratefully acknowledge support from the Center for Statistics and the Social Sciences at the University of Washington, the Donald R. Matthews Fund, and the Benificus Foundation. Data on state policy responses to COVID-19, including policies on masks, are available at http://covid19statepolicy.org.

## Footnotes

1 We use “waves” to denote sustained periods of surging COVID-19 cases across the United States which were followed by sustained periods of declining cases, while noting that some states followed different patterns of surging and declining new cases over the course of the epidemic (Zhang, Marioli, and Gao, 2021).

2 Mississippi has a particularly complex history of statewide and state-coordinated county level mandates. On 12 May 2020, Governor Tate Reeves issued a Level 2 mask mandate initially applying to just seven counties which accounted for a total of less than a quarter million residents or approximately 7% of Mississippi’s 2019 population (State of Mississippi, 2020*a*; U.S. Census Bureau, Population Division, 2020*a*). The list of counties placed under this mandate varied over the following months, until on 4 August 2020, Reeves issued a statewide mandate at Level 3 (State of Mississippi, 2020*b*), only to end that mandate on 30 September 2020 (State of Missis-sippi, 2020*c*). On 19 October 2020, Reeves again issued substate mask requirements, this time at Level 2 and initially applied to just eight counties comprising more than half a million residents (State of Mississippi, 2020*d*). By 23 December 2020, 78 of Mississippi’s 82 counties and more than 99% of the state population were covered by the substate mandate (State of Mississippi, 2020*e*; U.S. Census Bureau, Population Division, 2020*a*), which Reeves then ended on 3 March 2021 (State of Mississippi, 2021).

3 Population coverage of mask mandates calculated by the authors using 2019 population estimates from the US Census (U.S. Census Bureau, Population Division, 2020*b*).

4 We create daily measures of the trend in deaths for each state-say, we first construct a retrospective two-week window of the seven-day moving average of daily deaths per million population. (As in our measures of death rates, seven-day averages are again needed to smooth out idiosyncracies in reporting over different days of the week.) We then estimate the linear trend on this window, and use this slope estimate as our trend variable.

5 This control is available over the whole period only from the COVID Tracking Project.

6 While some cities and counties adopted mask mandates on their own initiative, such efforts are outside the scope of our analysis, which seeks to understand actions by state political leaders. For our purposes, the potentially relevant substate mask mandates are those substate mandates of at least Level 2 which were coordinated by the state government.

## Notes

### Competing Interest Statement

The authors have declared no competing interest.

### Summary of Updates

Data and analysis updated through 31 December 2020.

